# Delay Discounting, Nucleus Accumbens Activation, and Alcohol Use Trajectories in Young Adults with a Family History of Alcohol Use Disorder

**DOI:** 10.1101/2025.09.02.25334926

**Authors:** Amanda Elton, Jacqueline Aloumanis, JeeWon Cheong, J. Hunter Allen, Jill A. Star, Sara Jo Nixon

**Affiliations:** Department of Psychiatry, University of Florida, Gainesville, FL, 32610; Department of Neuroscience, University of Florida, Gainesville, FL, 32610; Center for Addiction Research & Education, University of Florida, Gainesville, FL, 32610; Department of Health Education & Behavior, University of Florida, Gainesville, FL, 32610; Department of Psychology and Neuroscience, University of North Carolina, Chapel Hill, NC, 27599

## Abstract

A family history (FH) of alcohol use disorder (AUD) is associated with increased personal risk for alcohol misuse and AUD. FH is also related to increased impulsivity as evidenced by performance on delay discounting tasks, representing a potential mechanistic link between FH and alcohol misuse. Delay discounting tasks assess individual differences in preferences for smaller, immediate versus larger, delayed rewards, the former being linked to substance misuse. Decision-making on such tasks is underpinned by multiple neural systems, including those supporting reward valuation, cognitive control, and future-oriented thinking. We hypothesized that FH would be associated with differences in one or more neural systems related to delay discounting, with these differences being related to increases in alcohol misuse in young adulthood. We tested 163 first-year college students (ages 18-19) with varying levels of familial risk for AUD on an fMRI delay discounting task. Alcohol misuse was self-reported at baseline and in three yearly follow-up surveys using the Alcohol Use Disorders Identification Test (AUDIT). Alcohol misuse was modeled using a latent growth model, and we examined mediation between FH and alcohol misuse trajectory (AUDIT intercept and slope) through functional activation of brain regions implicated in reward valuation (nucleus accumbens), cognitive control (middle frontal gyrus), and future-oriented thinking (hippocampus). FH was associated with greater activation of the nucleus accumbens, which in turn predicted a steeper AUDIT slope. No other mediators were significant. Our results demonstrate that nucleus accumbens function may be a key mechanism by which FH increases risk for alcohol misuse and AUD.

## INTRODUCTION

Alcohol use disorder (AUD) most frequently develops during late adolescence and young adulthood, affecting approximately 15% of individuals between the ages 18-25 in the United States[1]. One of the strongest predictors of developing AUD is a family history (FH) of the disorder. Familial AUD increases an individual’s risk for AUD by 3-4 fold[2,3] through both heritable and environmental effects[2]. Moreover, 22% of adults in the US having at least one biological parent with AUD[4]. Individuals with biological relatives affected by AUD are at elevated risk for earlier initiation as well as more severe trajectories of alcohol-related problems across the lifespan[5,6]. However, the neurocognitive pathways through which familial risk is transmitted remain incompletely understood.

One mechanism hypothesized to underlie both FH and addiction more generally, is delay discounting, or the tendency to discount delayed rewards, resulting in an exaggerated preference for immediate rewards. In laboratory delay discounting tasks, individuals choose between larger, delayed rewards and smaller, immediate (or less delayed) rewards to derive an individualized measure of impulsive decision-making. Delay discounting has been consistently associated with substance use severity and treatment outcomes[7,8]. Moreover, higher rates of delay discounting prior to alcohol initiation predict adolescent and young adult drinking trajectories[9], suggesting this form of decision-making relates to the development of alcohol misuse. Importantly, individuals with FH exhibit steeper discounting rates, suggesting an inherited or environmental vulnerability toward impulsive decision-making[10,11]. In fact, the effects of environmental and familial risk factors on delay discounting behavior have been linked to future substance use[12,13]. Thus, FH may relate to greater alcohol drinking trajectories through its effects on decision-making processes involving immediate versus delayed rewards. Specifically, the association of FH with impulsive decision-making and alcohol use may reflect functional differences in the brain regions that underlie intertemporal decision-making.

Existing data indicate that decisions involving choices between immediate and delayed rewards engage separable neural systems that support valuation, future-oriented thinking, and cognitive control. Reward valuation and motivation depend on nucleus accumbens (NAcc) activity[14,15], and greater NAcc activation is associated with making impulsive choices involving immediate rewards[16]. The hippocampus contributes to episodic representation, including episodic prospection, or imaging future events. This subcortical region is thought to reduce discounting of delayed rewards through its role in allowing individuals to imagine themselves in the future[17]. Prefrontal cortical regions are engaged during selection of larger, delayed rewards[18,19]. Activation of these regions support cognitive control, enabling individuals to inhibit impulsive tendencies. These key systems are each associated with variation across individuals, leading to individual differences in choice behavior. Individual differences related to FH may be driven by alterations in one or more of these systems that promote impulsive decision-making, ultimately increasing susceptibility to substance misuse and addiction.

Indeed, prior research suggests that FH is associated with altered function across these neural systems, even in substance-naïve youth. For example, adolescents with FH have shown increased striatal reactivity to reward cues[20], altered connectivity in frontostriatal networks[21], and larger NAcc volumes[22]. FH is also associated with greater engagement of prefrontal regions during cognitive control tasks among adolescents, suggesting less efficient processing in these regions[23]. Alcohol-naïve adolescents with a family history of AUD may also exhibit patterns of altered hippocampal volume[24], suggesting that hippocampal functioning may be impacted as well. However, findings on hippocampal volumes could also reflect the association of FH with childhood trauma[25], which affects the hippocampus[26]. Together, these findings suggest that familial risk for AUD is associated with early and persistent differences in brain regions supporting reward valuation and processing, episodic prospection, and cognitive control, indicating potential brain differences that could underpin the observed steeper discounting behavior in these individuals.

The current study aims to connect these prior associations by examining the delay discounting-related neural mechanisms linking FH to alcohol use trajectories in early adulthood, specifically during college years, a developmental period marked by increased independence and alcohol consumption. Participants completed an fMRI delay discounting task at baseline to probe activation in regions associated with reward (NAcc), prospection (hippocampus), and cognitive control (prefrontal cortex). We applied latent growth mediation models to test the hypothesis that neural responses in these regions mediated the association between FH and alcohol use trajectory over four years. By integrating longitudinal alcohol use assessments with task-based fMRI and a theory-driven analytic approach, this study aims to clarify the neural mechanisms by which familial risk for AUD contributes to the development of hazardous patterns of drinking in young adults.

## METHODS

The study design included a baseline fMRI scan, baseline surveys, and three yearly follow-up surveys completed online.

### Participants

165 first-year college students were recruited from multiple universities in the area surrounding Chapel Hill, North Carolina. Inclusion criteria were that participants were 18–19 years old and enrolled in their first year in a 4-year undergraduate degree program. Exclusion criteria were MRI contraindications, routine psychoactive medication or substance use other than non-disordered alcohol use, neurological disorders, and psychiatric disorders other than past mood or anxiety disorders. Psychiatric disorders were assessed with a Mini-International Neuropsychiatric Interview (M.I.N.I.) for DSM-IV[27], with DSM-5 criteria used to assess AUD and other substance use disorders. A five-panel urine drug screen was conducted prior to the scan, with no participants testing positive for cocaine, cannabis, opioids, amphetamines, or methamphetamine. An alcohol breathalyzer test was similarly conducted, with no positive results. Participants provided written informed consent prior to participation in this study, which was approved by the UNC Office of Human Research Ethics.

### Self-Report Data

Baseline surveys administered in REDCap assessed family history of AUD, childhood maltreatment histories, adolescent binge drinking frequency and recent alcohol use.

FH was assessed with the Family History Assessment Module, measuring likely AUD among first- and second-degree relatives. A family history density composite score by adding up a weighted total of the number of affected parents (0.5 for each), grandparents (0.25 for each), and maternal and paternal aunts and uncles (0.25/[total relatives in category] for each)[28].

The Childhood Trauma Questionnaire (CTQ)[29] was collected to assess childhood experiences of physical abuse, emotional abuse, sexual abuse, physical neglect, and emotional neglect, which were summed for a total score.

Adolescent binge drinking frequency was measured using a single question to assess the number of binge episodes prior to the age of 18: “Before the age of 18, how often did you have 5 or more drinks (4 or more if you are female) containing any kind of alcohol within a two-hour period?[30].”

The Alcohol Use Disorders Identification Test (AUDIT)[31] was administered at both baseline and in yearly follow-up surveys sent to participants’ emails via REDCap. Scores of 8 or greater are associated with harmful alcohol use.

Two participants were missing baseline survey data and were excluded from analyses. These same participants were missing fMRI data. Thus, the final sample included in analyses was 163 participants, with 148 contributing fMRI data.

Whereas baseline AUDIT data was available for all 163 participants, follow-up data was available for 128, 94, and 87 participants at the first, second, and third follow-ups respectively. AUDIT scores at baseline did not significantly relate to missingness at any timepoint, and AUDIT scores at the first and second follow-ups did not predict missingness at future follow-ups. No other variables tested – sex, FH, CTQ, adolescent binge drinking, and delay discounting behavior – were significantly related to missingness at any follow-up timepoint.

### Delay Discounting Task

A 48-trial pre-scan delay-discounting task using a rapid adjusting procedure was implemented outside the scanner to introduce participants to the task and to provide individualized starting reward values for the fMRI task at each delay[32]. The indifference points for each delay of the pre-scan task were also used to calculate a model-free, area-under-the-curve behavioral measure of delay discounting for $100 and $1000, at one week, one month, 6 months, or two years, where larger area-under-the-curve values reflect less discounting.

Participants next completed a 120-trial delay discounting task while undergoing fMRI, divided into two runs of 60 trials. Two hypothetical choices were presented on the left and right sides of the screen with options of a smaller monetary reward available “TODAY” or a larger monetary reward available at one week, one month, 6 months, or two years in the future. Larger rewards were either $100 or $1000, with the smaller reward adjusting based on prior choices to vary around an indifference point so that decision difficulty was relatively similar across participants. Each in-scanner trial began with a 1.0 second cue to indicate the trial type: “WANT,” “SOONER,” and “LARGER.” There were 14 LARGER trials and 14 SOONER trials, representing control trials in which participants were tasked with identifying the larger reward option or the reward available sooner, respectively. There were also 80 WANT trials in which participants selected their preferred reward. Cue text remained on the screen while the two monetary reward options were displayed for 5 additional seconds. Choices were made by a left or right button press on an MRI-compatible button box. Additionally, there were 12 null trials in which cues were presented without choices appearing to enable deconvolution of cue and decision-making trial components. Intertrial intervals ranged from 1-6 seconds. Trial types were pseudorandomly presented and balanced across runs.

Due to scanner or task malfunctioning, or incomplete scanning sessions, task fMRI data were unavailable or excluded for 17 subjects. Thus, 148 subjects had fMRI data available.

### MRI Data Acquisition

Blood oxygenation level-dependent (BOLD) fMRI data were collected with multiband echo-planar imaging (EPI) on a Siemens 3 T Prisma scanner with a 32-channel head coil.

The sequence included the following parameters: multiband factor=8, TR=800 ms, TE=37 ms, flip angle=52°, 2 mm isotropic voxels, 72 sagittal slices with interleaved acquisition, field of view (FOV)=208 × 208, bandwidth=2290 Hz/pixel. A mid-study scanner update led to a minor sequence adjustment for 27 subjects: bandwidth=2186 Hz/pixel and TE=38.2 ms. The first run of the delay discounting task was acquired with anterior-to-posterior (AP) phase encoding, whereas a posterior-to-anterior (PA) phase encoding was used for the second run. Each run lasted 8 minutes for a total scan time of 16 minutes.

A magnetization-prepared rapid gradient-echo (MPRAGE) T1-weighted image was also acquired to assist with registration and tissue segmentation: TR=2530 ms, TE=2.3 ms, flip angle=9°, 1 mm isotropic voxels, 176 sagittal slices, FOV=256 × 256.

### MRI data preprocessing

for structural T1-weighted (T1w) images and BOLD fMRI data were preprocessed using fMRIPrep[33]. Preprocessing of BOLD images included motion correction, slice-timing correction, susceptibility distortion correction, co-registration to anatomical images, spatial normalization, and estimation of confounding signals. Preprocessing for this study has been detailed previously[34].

### First-Level fMRI Analysis

We used a general linear modeling (GLM) approach (Friston) using 3dDeconvolve and 3dREML functions in AFNI (Cox) to evaluate the relative level of activation of brain regions across different trial types for each participant. Four trial types were modeled: all WANT trials with $100 as the delayed reward, all WANT trials with $1000 as the delayed reward, SOONER trials, and LARGER trials. Confounding signals related to 6 motion parameters, average WM and CSF time series, as well as the derivatives, squared values, and square of the derivatives for each of these measures were modeled as nuisance regressors. Additionally, the ten components derived using aCompCor from a combined mask combining WM and CSF voxels were also modeled[35]. To further reduce motion effects, we censored time points where the calculated framewise displacement was >0.5 mm.

The contrast of interest was the difference in estimated activation between WANT trials and control trials, e.g., (.5*WANT for $100 + .5*WANT for $1000) - (.5*SOONER + .5*LARGER). This contrast isolates brain activation related to subjective decision-making between smaller, immediate and larger, delayed rewards.

### Region-of-Interest (ROI) Approach

ROIs were selected from the Desikan-Killiany atlas based on neural processes known to underlie delay discounting behavior, specifically valuation, prospection, and cognitive control. In line with existing data-supported theory[36,37], we selected the bilateral nucleus accumbens to represent valuation, the bilateral hippocampus to represent prospection, and the bilateral caudal middle frontal gyrus to represent cognitive control (Figure 1). These regions closely map onto the implicated processes based on prior literature[16–19]. For each participant, their average contrast weight among voxels within each ROI for the WANT>Control contrast was calculated. Sample distributions of contrast values in these regions were examined, and any values greater than or less than three standard deviations from the mean were replaced with the value equal to three standard deviations from the mean to reduce the influence of extreme values.

**Figure 1.**
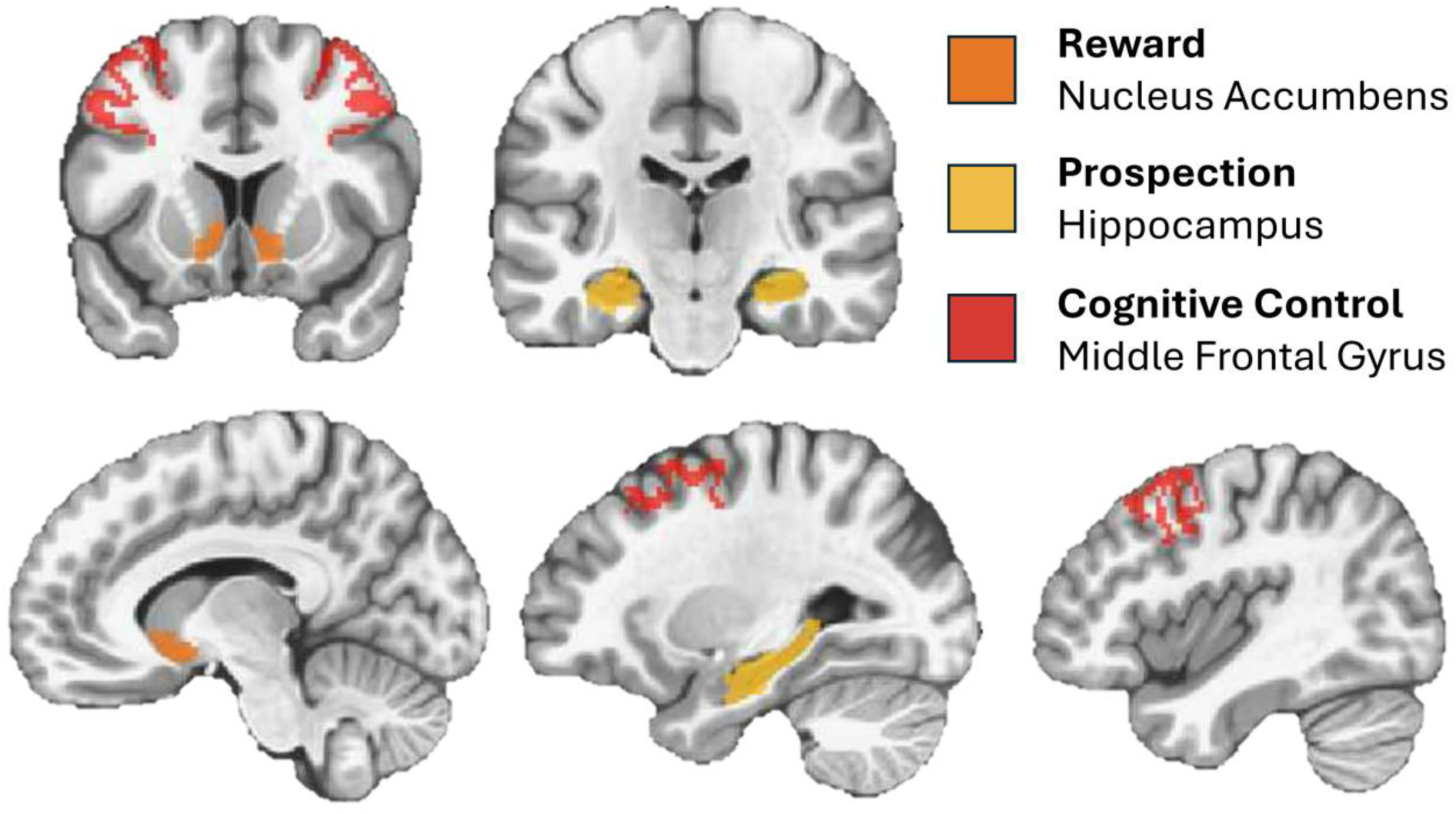
Regions-of-interest representing delay discounting neural processes. Nucleus accumbens activation represents reward valuation (orange). Hippocampal activation is involved in imagining the future or prospection (yellow). The middle frontal gyrus is involved in cognitive control (red). Regions were defined from the Desikan-Killiany atlas.

### Latent Growth Mediation Model

Structural Equation Modeling in Mplus 8.11[38] was utilized to estimate a Latent Growth Mediation Model (Figure 2) to examine the delay-discounting neural mechanisms that may link family history to alcohol use patterns. As previously described, the family history predictor variable was modeled using the weighted density score comprised of parents and second-degree relatives AUD-related behaviors. Alcohol use trajectory was modeled using AUDIT questionnaire scores collected annually over four years. The latent intercept specified to represent the baseline level of AUDIT scores, while the latent slope captured change per year across four years. To capture a primarily linear trajectory while allowing flexibility at the year 4 assessment, slope factor loadings were fixed to 0, 1, and 2, with the loading for the final timepoint freely estimated to improve model fit and facilitate model convergence. The model was estimated using MLR in Mplus, which yields robust standard errors under non-normality and handles missing data under the assumption of missing at random (MAR).

**Figure 2.**
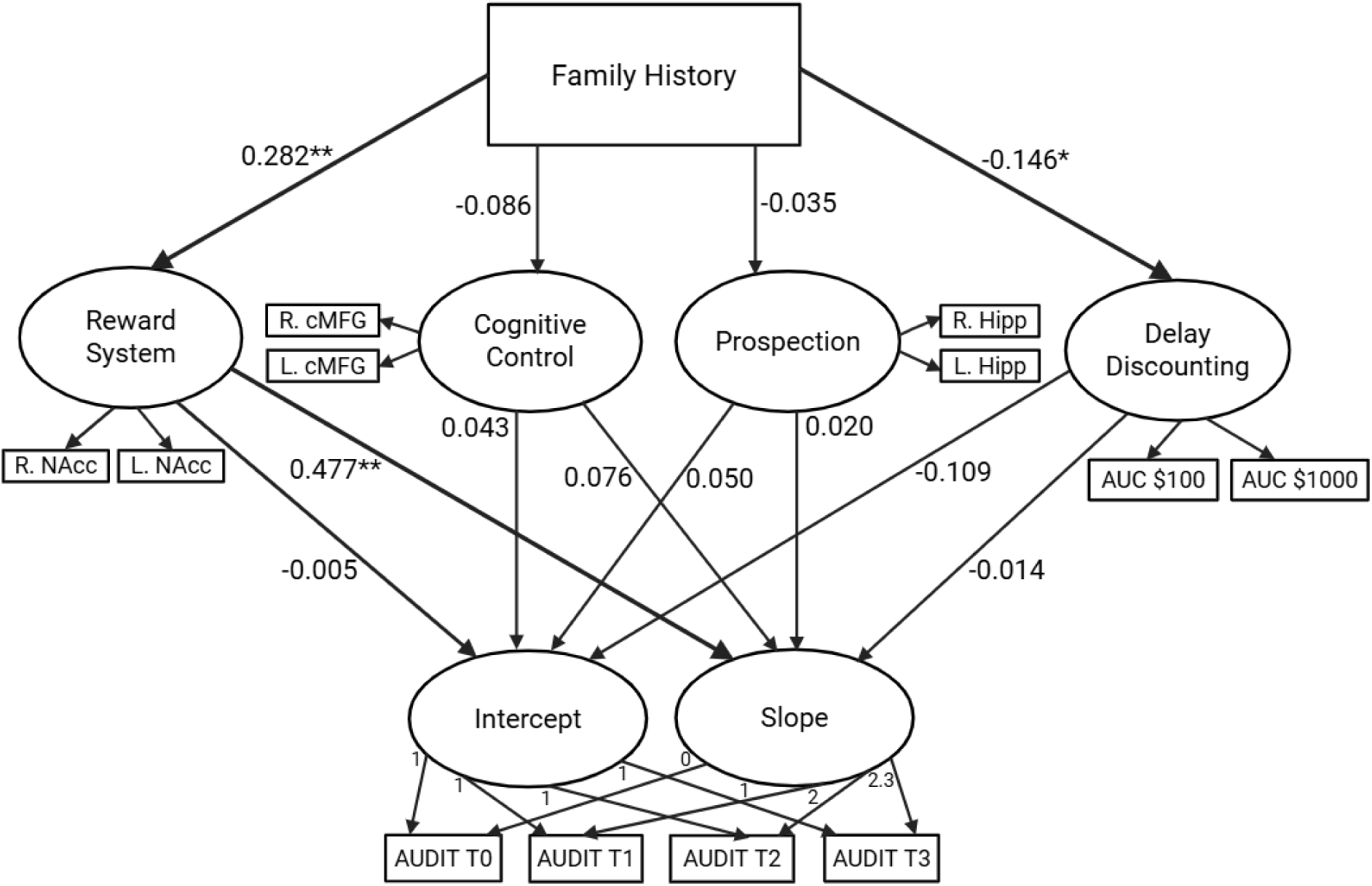
Latent growth mediation model of family history mediation of alcohol use trajectory through neural activation. * *p* < 0.05, ** *p* < 0.01, *** *p* < 0.001. AUC, area under the curve; AUDIT, Alcohol Use Disorders Identification Test; cMFG, caudal middle frontal gyrus; Hipp, hippocampus; NAcc, nucleus accumbens; L, left; R, right.

Participant sex was recorded at baseline with each participant identifying as male or female defined by their sex assigned at birth. Sex was used as a covariate through binary coding of female as 1 and male as 0. To control for higher rates of childhood abuse and neglect experienced by individuals with FH[25], we included total scores from the childhood trauma questionnaire[29] as a covariate. We controlled for adolescent heavy alcohol use with a binary variable indicating whether participants reported engaging in binge drinking (1+ episode) before age 18 (1=yes, 0=no).

Additionally, 12 subjects demonstrated no selection of immediate choices throughout the task, suggesting that the task failed to provide the same level of decision difficulty for this subgroup compared to other participants. Therefore, a covariate (1=no choice variation or 0=choice variation) was created to account for possible effects on fMRI activation.

To examine mediation (Figure 2), reward, prospection, and cognitive control were modeled as latent variables based on ROI activation in the NAcc, hippocampus, and caudal middle frontal gyrus, respectively, and were included as mediator variables. Additionally, task-related delay discounting performance (AUC) was included as a behavioral mediator, represented by a latent variable indicated by the $100 and $1000 options. The indirect path from family history to the latent slope through each mediator was calculated.

## RESULTS

Participant self-report and behavioral data are presented in Table 1.

**Table 1.** Descriptive statistics (means and standard deviations) for self-report and behavioral data.

| Variable | Mean | Std. Dev. | Median | Min | Max |
| --- | --- | --- | --- | --- | --- |
| Sex, n (%) | 105 (64%) |  |  |  |  |
| Binge drinking prior to age 18, n (%) | 41 (25%) |  |  |  |  |
| Childhood Trauma Questionnaire | 36.0 | 10.8 | 33 | 25 | 91 |
| Family History Density | 0.41 | 0.42 | 0.29 | 0.0 | 1.6 |
| AUDIT Baseline | 2.7 | 2.9 | 2 | 0 | 12 |
| AUDIT Follow-up 1 | 3.6 | 3.7 | 3 | 0 | 17 |
| AUDIT Follow-up 2 | 4.0 | 3.8 | 3 | 0 | 22 |
| AUDIT Follow-up 3 | 4.2 | 3.6 | 3 | 0 | 18 |
| Delay Discounting AUC \$100 | 55.0 | 27.8 | 52.2 | 2.6 | 98.4 |
| Delay Discounting AUC \$1000 | 612 | 283 | 606 | 36 | 984 |

AUDIT, Alcohol Use Disorders Identification Test; AUC area under the curve

The unconditional latent growth model to characterize the AUDIT trajectory shape indicated that mean AUDIT scores began at 2.719, with significant variability in baseline levels (Var=5.821, SE=1.311, p < .001). The average slope was positive (M=0.751, SE=0.169, p < .001), suggesting that AUDIT scores increased over time, with significant individual differences in rates of change (Var=1.452, SE=0.531, p=.006). The freely estimated slope loading for the final time point was 2.436, indicating that the increase in AUDIT scores from year 3 to year 4 was smaller than expected under strict linear growth, consistent with a leveling-off of alcohol use in the later period.

A latent growth mediation model of delay discounting-related neural mechanisms linking family history to patterns of alcohol use in early adulthood (Figure 2) demonstrated an acceptable model fit: X^2^(73)=78.211, *p*=0.440; CFI=0.998; TLI=0.997; RMSEA=0.010 [90% CI: 0.000, 0.046]; SRMR=0.040. All the factor loadings on the mediator variables were statistically significant (p < 0.001), indicating a strong measurement of the latent constructs.

Standardized coefficients of family history predicting neural mediators (Table 2) showed that family history significantly predicted reward-related neural activation in the nucleus accumbens (β=0.282, SE=0.103, p=0.006). Additionally, family history predicted lower area under the curve, i.e., more impulsive delay discounting behavior (β=−0.146, SE=0.070, p=0.037). Family history was not significantly associated with neural mediators representing prospection (β=−0.035, SE=0.102, p=0.734) or cognitive control (β=−0.086, SE=0.098, p=0.382). However, there was a noteworthy effect of CTQ scores on reduced activation in the neural representation of prospection (hippocampus) (β=−0.237, SE=0.093, p=0.010).

**Table 2.** Standardized model results for effects of family history and covariates on latent mediators.

| Predictors | Mediators |  |  |
| --- | --- | --- | --- |
|  | Reward |  |  |
| | $\beta$ | SE | p-value |
| Family History | 0.282 | 0.103 | 0.006 |
| Sex | -0.007 | -0.167 | 0.868 |
| Adolescent Binge Drinking | -0.053 | 0.079 | 0.504 |
| Childhood Trauma | 0.000 | 0.003 | 0.885 |
| No Choice Variation | 0.131 | 0.081 | 0.110 |
|  | Cognitive Control |  |  |
| | $\beta$ | SE | p-value |
| Family History | -0.086 | 0.098 | 0.382 |
| Sex | 0.027 | 0.083 | 0.747 |
| Adolescent Binge Drinking | 0.013 | 0.074 | 0.857 |
| Childhood Trauma | 0.002 | 0.776 | 0.438 |
| No Choice Variation | -0.144 | 0.160 | 0.368 |
|  | Prospection |  |  |
| | $\beta$ | SE | p-value |
| Family History | -0.035 | 0.102 | 0.734 |
| Sex | 0.047 | 0.031 | 0.127 |
| Adolescent Binge Drinking | -0.062 | 0.033 | 0.065 |
| Childhood Trauma | -0.004 | 0.001 | 0.010 |
| No Choice Variation | 0.040 | 0.057 | 0.486 |
|  | Delay Discounting |  |  |
| | $\beta$ | SE | p-value |
| Family History | -0.146 | 0.070 | 0.037 |
| Sex | 0.119 | 0.037 | 0.001 |
| Adolescent Binge Drinking | -0.033 | 0.043 | 0.440 |
| Childhood Trauma | 0.001 | 0.002 | 0.550 |
| No Choice Variation | 0.403 | 0.037 | 0.000 |

Latent variable for reward was comprised of the bilateral nucleus accumbens, cognitive control was comprised of the bilateral caudal middle frontal gyrus, prospection was comprised of the bilateral hippocampus, and delay discounting was comprised of the area-under-the-curve behavior for \$100 and \$1000 options.

The slope of alcohol use trajectory was significantly predicted by the reward (NAcc) mediator variable (β=0.477, SE=0.163, p=0.003). More detailed standardized model results for the latent intercept and slope can be found in Table 3.

**Table 3.** Standardized model results for effects of predictor variables and covariates on latent intercept and slope.

| Predictors | AUDIT Intercept |  |  |
| --- | --- | --- | --- |
| | $\beta$ | SE | p-value |
| Reward | 0.005 | 0.165 | 0.978 |
| Cognitive Control | 0.043 | 0.135 | 0.751 |
| Prospection | 0.050 | 0.131 | 0.701 |
| Delay Discounting | -0.109 | 0.104 | 0.293 |
| Family History | 0.130 | 0.102 | 0.205 |
| Sex | 0.096 | 0.081 | 0.234 |
| Adolescent Binge Drinking | 0.672 | 0.063 | 0.000 |
| Childhood Trauma | 0.014 | 0.181 | 0.857 |
|  | AUDIT Slope |  |  |
| | $\beta$ | SE | p-value |
| Reward | 0.477 | 0.163 | 0.003 |
| Cognitive Control | 0.076 | 0.179 | 0.670 |
| Prospection | 0.020 | 0.125 | 0.901 |
| Delay Discounting | -0.014 | 0.140 | 0.923 |
| Family History | -0.174 | 0.122 | 0.155 |
| Sex | -0.026 | -0.202 | 0.840 |
| Adolescent Binge Drinking | -0.174 | 0.107 | 0.103 |
| Childhood Trauma | 0.154 | 0.147 | 0.293 |

There was a significant indirect effect of FH on AUDIT slope through the reward (NAcc) latent mediator (β =0.321, SE= 0.155, p=0.039).

## DISCUSSION

This study yielded several novel findings. First, we identified the effects of FH – a major risk factor for AUD – on the neural correlates of delay discounting, revealing significantly elevated activation in the NAcc. Secondly, although the role of delay discounting as a mechanism risk for addiction has long been appreciated, studies identifying the neural mechanisms linking this vulnerability to substance misuse have been limited. In the current study, we found that NAcc activation during delay discounting prospectively predicted future alcohol use. Finally, using mediation analysis in a structural equation modeling framework, we identified the neural mechanisms linking FH to patterns of alcohol use in young adulthood. Specifically, greater activation of the NAcc during decision-making for immediate versus delayed rewards mediated the association between FH and steeper alcohol use trajectories in young adulthood. These results support theoretical models of the nucleus accumbens and reward circuit functioning as a neurobiological mechanism of risk for addiction[39,40], and indicate this may be a major neurobiological pathway through which FH promotes alcohol misuse and AUD.

The NAcc, a core region of the mesolimbic dopamine system, has been implicated in reward valuation and the motivational salience of appetitive cues[39,41]. Increased activity in this region is observed in individuals with substance use disorders in response to conditioned drug cues[42]. Supporting its possible role in promoting risk for addiction, alterations in NAcc structure and task-free fMRI measures of NAcc function have also been observed in youth with familial risk for AUD prior to their initiation of substance use[21,22]. Other studies have demonstrated blunted NAcc activation to reward-predicting cues in monetary incentive delay tasks among youth with a family history of AUD[20,43]. The current findings add to this growing evidence of effects of FH on NAcc reward function and expand this observation to intertemporal decision-making for rewards, suggesting a more generalized deficit in reward-related functioning.

Investigations of the effects of FH on neural correlates of delay discounting have thus far been limited. A prior study of 125 adolescents with a family history of substance use disorders found no effects of FH on fMRI activation during a delay-discounting task despite the detection of behavioral differences[44]. However, that study included whole-brain analyses, with no clusters surviving correction for multiple comparison, as well as underpowered main effects of task in the striatum, suggesting the discrepancy could stem from differences in methodology and/or power. Another fMRI study found that substance-naïve youth with FH (n=35) had greater activation in the posterior insula, thalamus, and parahippocampal gyrus compared with controls (n=24)[45]. A study of 33 adolescents linked FH to changes in white matter structure, which related to slower response times during a delay discounting task[11]. Results from the current study yielded significant effects of FH based on our ROI approach, and further suggest these effects are a neural mechanism through which FH promotes future drinking behavior.

While prior studies have shown steeper delay discounting in individuals with a family history of AUD[46], our study identifies the underlying neural activation patterns associated with this behavioral expression and links it to longitudinal alcohol use trajectories. The fact that this mediation occurred via neural, rather than behavioral markers suggests added explanatory power of neuroimaging in identifying mechanisms of risk. In fact, although FH was associated with steeper behavioral discounting in the current sample, this behavior did not significantly mediate effects on drinking. Although discounting behavior is observable and predictive, it is likely that different patterns of activation across multiple brain systems can lead to similar behaviors due to their relative roles in supporting immediate versus delayed choices. Thus, NAcc function may be a more predictive marker of risk for increased alcohol misuse in young adults with FH than delay discounting behavior.

Although we also tested candidate mediators related to prospection (hippocampus) and cognitive control (prefrontal cortex), NAcc was the only significant neural predictor of alcohol use trajectories in this college sample. It is possible that other brain systems involved in delay discounting would be more predictive of alcohol use in other contexts or age groups. For example, in a sample of adolescents (ages 12-18) in an outpatient treatment program, substance use during the treatment period was related to neural activation during delay discounting in a medial temporal lobe “limbic” network (greater activation related to greater substance use), but not a network centered in the nucleus accumbens[47]. Network functional connectivity in the default-mode network, which supports future-oriented thinking, related to longer-term, post-treatment substance use. Qualitative differences between these studies suggest the neural processes supporting treatment response may differ from those in a non-treatment-seeking college sample.

Another longitudinal study using IMAGEN data identified correlational trends for lower AUDIT scores at ages 16 and 18 in participants with greater fMRI activation in the dorsolateral prefrontal cortex during delay discounting at those time points, but there was no significant association of NAcc with alcohol use when examined as an ROI. The role of prefrontal regions in alcohol misuse in adolescence may stem from high interindividual variation due to in rapid development of cognitive control during that period[48], whereas the NAcc may play a larger role in young adulthood. Overall, the current findings suggest reward sensitivity may represent a proximal neurobiological link between family history and young adult drinking behavior. Other cognitive processes may exert their influence in other developmental periods, in other stages of AUD development (e.g., initiation or transition to compulsive use), or in different contexts (e.g., during abstinence or treatment).

The findings of this study have implications for early intervention among young adults. Reducing hazardous alcohol use, especially on college campuses, remains an important goal of intervention research. Although current NIAAA recommendations for individual-level strategies for college students include some that target high-risk student groups (e.g., Greek organizations), none currently utilize a precision-medicine approach based on neurocognitive features of the at-risk individual. Thus, an improved understanding of the individual-level factors that promote alcohol misuse and AUD among young adults, including their neural and behavioral mechanisms, represents a crucial step toward reducing the personal and economic burden of hazardous drinking. The current findings suggest that young adults with heightened reward-related reactivity might benefit from targeted strategies designed to modulate reward sensitivity or enhance the salience of non-drug rewards. In fact, the mesolimbic dopamine system, with the ventral tegmental area as a key hub, has been implicated in the ability of natural rewards to protect against the development of AUD[49]. Given the association of NAcc activation with familial risk, such interventions may be particularly effective in young adults with a family history of AUD.

Some study limitations should be noted. First, alcohol use may be driven by different neural mechanisms in other age groups or contexts, which would limit the generalizability of these findings. Second, we only focused on theory-derived brain regions, but it is also possible that FH promotes alcohol use through other brain regions not included in the analysis. Third, it is possible that there are sex differences in the role of FH and the brain in alcohol drinking that this study was not powered to examine[34].

Given the high prevalence of FH[4] and the negative outcomes associated with this risk factor[5,6], the current study sought to uncover the neurobiological mechanisms by which FH increases risk for AUD. The findings contribute to mounting evidence that the NAcc is altered in individuals with high familial risk for AUD and suggest that differences in reward circuitry function confer vulnerability for AUD by biasing individuals toward immediate rewards. The study offers novel insight into the brain mechanisms of familial risk for AUD and highlights the reward system as a potential neural target of intervention in at-risk individuals.

## Data Availability Statement

The data can be made available upon request to Amanda Elton.

## Acknowledgements

AE would like to posthumously acknowledge the contributions of Charlotte Ann Boettiger, PhD and thank her for her mentorship on this project.

## Author Contributions

AE – Conceptualization, Investigation, Formal analysis, Writing – original draft; JA Visualization, Writing – original draft; JWC – Methodology, Writing – review & editing; JHA

Investigation, Writing – review & editing; JAS – Investigation; SJN – Supervision, Writing – review & editing

